# Clinical and pathological characteristics of 2019 novel coronavirus disease (COVID-19): a systematic reviews

**DOI:** 10.1101/2020.02.20.20025601

**Authors:** Yaqian Mao, Wei Lin, Junping Wen, Gang Chen

## Abstract

**Importance:** In 2002-2003, a severe pulmonary infectious disease occurred in guangdong, China. The disease was caused by severe acute respiratory syndrome coronavirus (SARS-CoV), 17 years apart, also happen in China, and also a novel coronavirus (SARS-CoV-2), this epidemic has posed a significant hazard to people’s health both China and the whole world.

**Objective:** Summarized the latest epidemiological changes, clinical manifestations, auxiliary examination and pathological characteristics of COVID-19.

**Evidence Review:** PubMed database were searched from 2019 to 2020 using the index terms “novel coronavirus” or “COVID-19” or “2019-nCoV” or “SARS-CoV-2” and synonyms. Articles that reported clinical characteristics, laboratory results, imageological diagnosis and pathologic condition were included and were retrospectively reviewed for these cases. This paper adopts the method of descriptive statistics.

**Results:** 34 COVID-19-related articles were eligible for this systematic review,Four of the articles were related to pathology. We found that Fever (86.0%), cough (63.9%) and Malaise/Fatigue (34.7%) were the most common symptoms in COVID-19. But in general, the clinical symptoms and signs of COVID-19 were not obvious. Compared with SARS, COVID-19 was transmitted in a more diverse way. The mortality rates of COVID-19 were 2.5%, and the overall infection rate of healthcare worker of COVID-19 was 3.9%. We also found that the pathological features of COVID-19 have greatly similar with SARS, which manifested as ARDS. But the latest pathological examination of COVID-19 revealed the obvious mucinous secretions in the lungs.

**Interpretation:** The clinical and pathological characteristics of SARS and COVID-19 in China are very similar, but also difference. The latest finds of pathological examination on COVID-19 may upend existing treatment schemes, so the early recognition of disease by healthcare worker is very important.

**Key Points:** *Question:* What can we learn from the clinical manifestations and pathological features of 2019 novel coronavirus disease (COVID-19)?

*Findings:* In this review, we found COVID-19 was transmitted in a more diverse way than Severe acute respiratory syndrome (SARS). Fever, cough and Malaise/Fatigue were the most common symptoms. We also found that the SARS-CoV-2 has the same cell entry receptor ACE2 as SARS-CoV, and they have similar pathological mechanisms like Acute respiratory distress syndrome (ARDS).

*Meaning:* This review aims to give people a more comprehensive understanding of COVID-19 and to continuously improve the level of prevention, control, diagnosis and treatment.

## Introduction

In December 2019, a group of unidentified pneumonia patients was found to be linked to a seafood wholesale market in wuhan, China^[1]^. By analyzing samples from pneumonia patients, we identified a previously unknown subcoronavirus, which we named 2019-novel coronavirus (2019-nCoV). WHO has officially named the virus severe acute respiratory syndrome coronavirus 2 (SARS-CoV-2) and the disease corona virus disease 2019 (COVID-19)^[2]^. By Mar 8, 2020, more than 105586 cases of COVID-19 pneumonia had been reported in countries and regions around the world, the cumulative death toll stands at 3584^[3]^. The United States, Australia, Germany, Vietnam, South Korea, Canada, Brazil, Nepal and Japan over 102 countries are also seeing increasing Numbers of travel-related cases and human-to-human transmission^[3-11]^. COVID-19 is transmitted primarily through droplets and contact, recent studies^[4, 12]^ showed that SARS-CoV-2 was detected in stool samples of patients, which makes it possible for gastrointestinal transmission.

Seventeen years ago, a severe acute respiratory syndrome (SARS) happened in Guangdong Province, China, and soon spread to other countries and regions^[13-16]^. By the end of the outbreak, 26 countries had reported 8,098 possible SARS cases and 774 deaths^[14]^. SARS-CoV and SARS-CoV-2 are both enveloped RNA viruses and widely distributed in humans, other mammals and birds. Recent studies analysis showed that the novel coronavirus had 78% nucleotide homology with human SARS–CoV^[17, 18]^. As. more and more COVID-19 cases are diagnosed, we find that SARS and COVID-19 share many similarities in clinical manifestations, epidemiology, and mode of transmission. At present, while the epidemic in China is gradually under control, Europe and the Middle East have shown a rapid spread of the epidemic. For example, the number of infections in South Korea, Italy, Iran and other countries has soared recently^[3]^. So far, our understanding of COVID-19 remained to be limited, Especially in pathology. This report summarizes the initial clinical description and Pathological discovery of SARS and COVID-19 in different countries. We also compared the pathological features of SARS-CoV and SARS-CoV-2 to provide a reference for the diagnosis and treatment of COVID-19 by describing and emphasizing the characteristic changes of SARS-CoV.

## Methods

This study aimed to summary the case of COVID-19 on clinical features, epidemiological findings, laboratory and imageological examination, pathological studies and treatment options in different countries.

### Search strategy

This study was a review that was conducted in 2020. Searches were done in scientific databases of PubMed, based on the combination of related keywords based of Mesh terms **(Table 1)**. One researcher (Y.Q.M), a professional clinician searched PubMed for all published articles regarding COVID-19 up to March 9, 2020. A second researcher (W. L), a professional clinician with expertise in systematic reviews, repeated the first reviewer’s search independently. Both searches agreed completely. All steps of searches were done based on the Preferred Reporting Items for Systematic Reviews and Meta-Analyses (PRISMA) checklist. The searches were limited to papers published in the English and Chinese language. Institutional review board approval and informed consent were not obtained given that the study was a systematic review of the literature, we limited our study to published information and did not engage with any human subjects.

**Table 1.** Search strategy of the research

| Search strategy |  |
| --- | --- |
| Database | PubMed |
| Limits | Language (in English or Chinese), Species (studies on humans) |
| Data | 2019 to March 9, 2020 |
| #1 (MeSH) | severe acute respiratory syndrome coronavirus 2 |
| #2 (Entry Terms) | “Wuhan coronavirus” or “Wuhan seafood market pneumonia virus”<br>or “COVID 19 virus” or “COVID-19 virus” or “coronavirus disease<br>2019 virus” or “SARS-CoV-2” or “SARS 2” or “2019-nCoV” or<br>“2019 novel coronavirus” |
| Search | #1 or #2 |

### Inclusion and exclusion criteria

The inclusion criteria were original articles, retrospective case series and case report of COVID-19 infection, including clinical features, epidemiological findings, laboratory and imageological examination, treatment options or pathological studies. Exclusion criteria were unavailable full text, no target observations and other article types, Other article types included review articles, comment, news, etc. Also conference articles were excluded. The data extraction of categories, including the author’s name, publication year, countries, age, sex, clinical features, epidemiological findings, laboratory and imageological examination, treatment options, pathological studies, etc. After data extraction, we summarized and reported the findings in tables and figures according to the objectives of the study. Two researchers (Y.Q.M and W.L who were specialist physicians) reviewed each of these articles in detail. Each researcher identified every article that concerned clinical characteristics or pathologic studies of COVID-19 infection. These search results are submitted to third parties (J.W.P who was a professionally trained physician), who review discrepancies and make decisions in the event of disagreement.

### Statistical analysis

Categorical variables were summarized as frequencies and percentages, and continuous variables were described using a range, median and interquartile ranges (IQR) values, Mean and standard deviation (SD).

## Results

As of March 9, 2020, we identified 34 studies, Among them, 33 articles^[2, 4-11,19-42]^ come from the PubMed database, and one article^[43]^ about pathology comes from a Chinese journal **(Figure 1)**. Three of the articles were related to pathology^[41-43]^. The characteristics of 34 studies are shown in **Table 2**. Articles on the characteristics of COVID-19 mainly came from China, as well as from countries with a small number of current case reports, such as the United States, Germany, Australia, Vietnam, Thailand, Japan, Canada, Brazil, Nepal and South Korea.

**Table 2:** Summary the characteristics of 34 studies that analysed the clinical or pathological characteristics of COVID-19 in different countries.

| ID<sup>1</sup> | Study | Year | Country | SS<sup>2</sup><br>(n) | Age (days,<br>months and years) | Sex/Male<br>(n, %) | Healthcare<br>worker (n, %) | Severe<br>patients (n, %) | Death<br>toll(n, %) | Mode of Transmission |
| --- | --- | --- | --- | --- | --- | --- | --- | --- | --- | --- |
| 1 | Kim, Jin Yong. | 2020 | Korea<sup>[8]</sup> | 1 | 35 | 0 | 0 | 0 | 0 | human-to-human |
| 2 | Holshue, M. L. | 2020 | United States<sup>[4]</sup> | 1 | 35 | 1(100.0) | 0 | 0 | 0 | human-to-human, fecal-oral route ? |
| 3 | Phan, L. T. | 2020 | Vietnam<sup>[6]</sup> | 2 | 27, 65 | 2(100.0) | 0 | 0 | 0 | human-to-human, familial cluster |
| 4 | Rothe, C. | 2020 | Germany<sup>[7]</sup> | 1 | 33 | 1(100.0) | 0 | 0 | 0 | human-to-human, asymptomatic transmission |
| 5 | N I R S T<sup>4</sup> | 2020 | Australia<sup>[5]</sup> | 12 | 21-66 | 7(58.3) | 0 | 1(8.3) | 0 | human-to-human |
| 6 | Pongpirul,W.A. | 2020 | Thailand<sup>[19]</sup> | 1 | 51 | 1(100.0) | 0 | 0 | 0 | human-to-human |
| 7 | Arashiro,Takeshi | 2020 | Japan<sup>[20]</sup> | 2 | 27, 35 | 1(50.0) | 0 | 0 | 0 | human-to-human |
| 8 | Rodriguez-MoralesAJ | 2020 | Brazil<sup>[9]</sup> | 1 | 61 | 1(100.0) | 0 | 0 | 0 | human-to-human |
| 9 | Shrestha R | 2020 | Nepal<sup>[10]</sup> | 1 | 32 | 1(100.0) | 0 | 0 | 0 | human-to-human |
| 10 | Silverstein WK | 2020 | Canada<sup>[11]</sup> | 1 | 56 | 1(100.0) | 0 | 0 | 0 | close contact |
| 11 | Wang, D. | 2020 | China<sup>[21]</sup> | 138 | 22-92 | 75(54.3) | 40(29.0) | 36(26.1) | 6(4.3) | human-to-human, hospital-related |
| 12 | Chen, Nanshan | 2020 | China<sup>[22]</sup> | 99 | 21-82 | 67(67.7) | 0 | 23(23.2) | 11(11.1) | human-to-human, close contact |
| 13 | Guan, W. J. | 2020 | China<sup>[23]</sup> | 1099 | 47(35-58)* | 637/1096<br>(58.12%) | 38(3.5%) | 173(15.7) | 15(1.4) | human-to-human, family cluster,<br>fecal-oral route? |
| 14 | Huang, C. | 2020 | China<sup>[24]</sup> | 41 | 49(41-58) | 30(73.2) | 0 | 13(31.7) | 6(14.6) | human-to-human, family cluster, close contact |
| 15 | Chen, Huijun | 2020 | China<sup>[2]</sup> | 9 | 26-40 | 0 | 0 | 0 | 0 | close contact, familial cluster |
| 16 | China CDC<sup>5</sup> | 2020 | China<sup>[25]</sup> | 44672 | 30-79 (86.6)<sup>#</sup> | 22981(51.4) | 1716(3.8) | 8255(18.5) | 1023 (2.3) | human-to-human, close contact, familial cluster |
| 17 | Shi H | 2020 | China<sup>[26]</sup> | 81 | 49.5±11 | 42(51.9) | 15(18.5) | NA<sup>3</sup> | 3(3.7) | human-to-human, close contact, familial cluster |
| 18 | Li Q | 2020 | China<sup>[27]</sup> | 425 | 59(15-89) | 240(56.5) | 15(3.5) | NA | NA | human-to-human, close contact |
| 19 | Song, F. | 2020 | China<sup>[28]</sup> | 51 | 16-76 | 25(49.0) | NA | NA | NA | close contact |
| 20 | Chen, L. | 2020 | China<sup>[29]</sup> | 29 | 56(26-79) | 21(72.4) | 0 | 14(48.3) | 2(6.9) | close contact |
| 21 | Zhou, Fei | 2020 | China<sup>[30]</sup> | 191 | 18-87 | 119(62.3) | NA | 119(62.3%) | 54(28.3) | human-to-human, close contact |
| 22 | Xu XW | 2020 | China<sup>[31]</sup> | 62 | 41(32-52) | 35(56) | 0 | 1(1.6) | 0 | human-to-human, familial cluster |
| 23 | Yang, W. | 2020 | China<sup>[32]</sup> | 149 | 45.11±13.35 | 81(54.4) | 0 | 0 | 0 | human-to-human, close contact |
| 24 | Xu YH | 2020 | China<sup>[33]</sup> | 50 | 3-85 | 29(58.0) | 0 | 13(26.0%) | NA | human-to-human, close contact |
| 25 | Wang, D | 2020 | China<sup>[34]</sup> | 31 | 7 yr and 1 mo<br>(6 mo -17 yr) | 15(48.4) | 0 | 0 | 0 | close contact, familial cluster |
| 26 | Liu K | 2020 | China<sup>[35]</sup> | 137 | 57(20-83) | 61(44.5) | 0 | NA | 16(11.7) | human-to-human |
| 27 | Zhang JJ | 2020 | China<sup>[36]</sup> | 140 | 57(25-87) | 71(50.7) | 3(2.1) | 58(41.4) | NA | human-to-human, familial cluster |
| 28 | Zhao W | 2020 | China<sup>[37]</sup> | 101 | 17-75 | 56(55.4) | NA | 14(13.9) | NA | human-to-human, familial cluster |
| 29 | Yang, Xiaobo | 2020 | China<sup>[38]</sup> | 52 | 59.7±13.3 | 35(67.3) | NA | 52(100) | 32(61.5) | human-to-human |
| 30 | Tian, Sijia | 2020 | China<sup>[39]</sup> | 262 | 47.5(1-94) | 127(48.5) | 0 | 46(17.6) | 3(1.1) | human-to-human, close contact, familial cluster |
| 31 | Xu, Xi | 2020 | China<sup>[40]</sup> | 90 | 50(18-86) | 39(43.3%) | NA | NA | NA | human-to-human, close contact |
| 32 | Tian, S. | 2020 | China<sup>[41]</sup> | 2 | 73, 84 | 1(50.0) | 0 | 1(50.0) | 1(50.0) | human-to-human |
| 33 | Xu, Zhe | 2020 | China<sup>[42]</sup> | 1 | 50 | 1(100.0) | 0 | 1(100.0) | 1(100.0) | human-to-human |
| 34 | Liu qian | 2020 | China<sup>[43]</sup> | 1 | 85 | 1(100.0) | 0 | 1(100.0) | 1(100.0) | NA |

COVID-19:2019 novel coronavirus disease; Data are median (IQR), mean (SD), n (%), n/N (%), rang; <sup>1</sup>Study ID: indicate the 1st to 34th study; <sup>2</sup>SS:Sample Size; <sup>3</sup>NA :not applicable; <sup>4</sup>N I R S T :National Incident Room Surveillance Team; <sup>5</sup>CDC: Centers for Disease Control and Prevention; \*: indicate the the whole spectrum of age; <sup>#</sup>:indicate 86.6 % of the patients were between the ages of 30 and 79.

**Figure 1.**
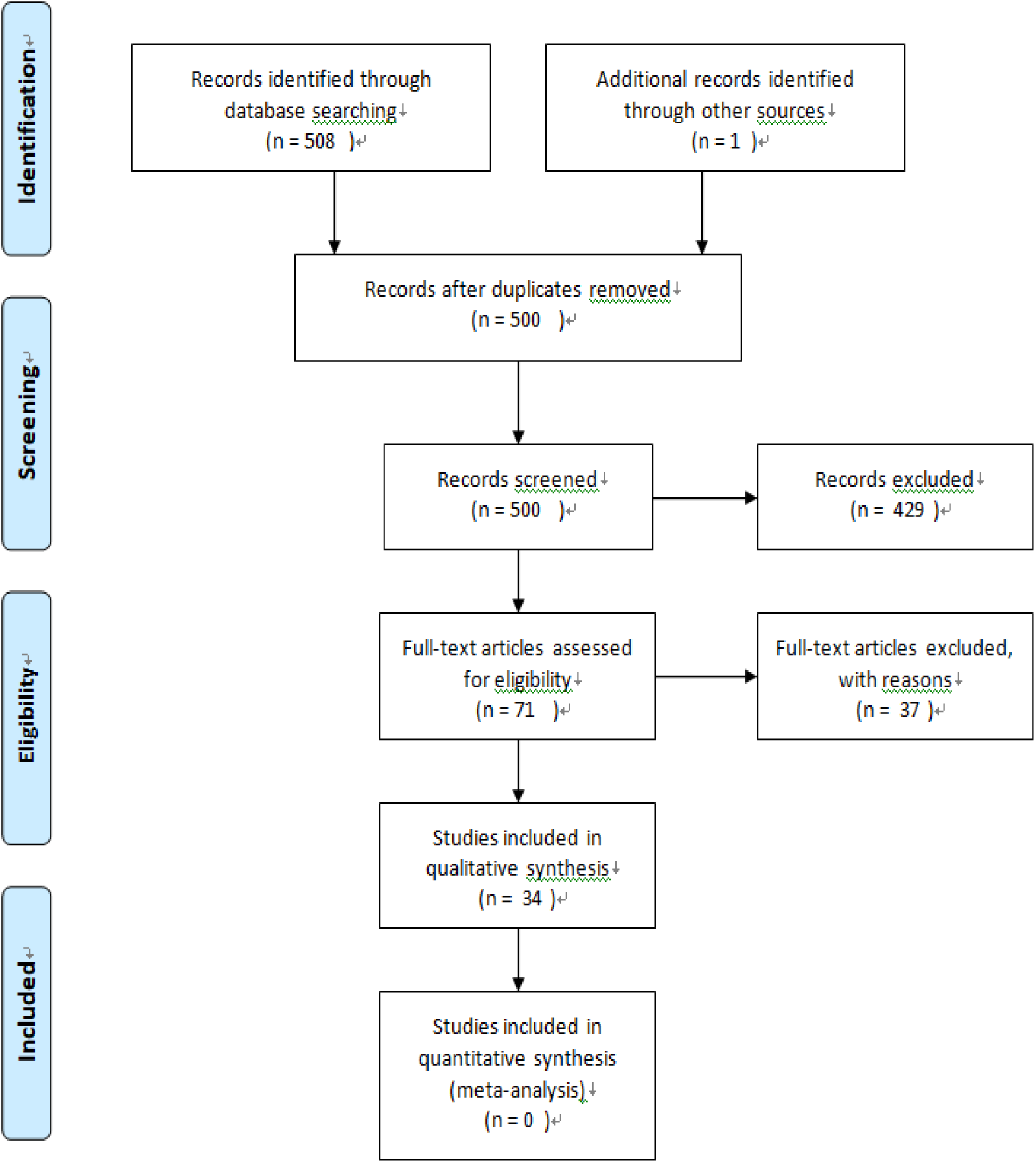
Flow Diagram for Selection of Studies.

### Demographic and clinical characteristics

Demographic and clinical characteristics are shown in **Table 3**. In COVID-19, we found 3.9% were health workers, and 51.7 % were male (male: female=1.07:1). The most common clinical manifestation of COVID-19 were Fever (86.0%), Cough (63.9%), Malaise/Fatigue/Confusion (34.7%), Sputum production (28.9%), Shortness of breath (19.7%) and Myalgia (18.8%), whereas diarrhea (5.7%) and Nausea/vomiting (6.1%) were rare. 26.7% of patients in COVID-19 had at least one underlying disorder (i.e., hypertension, diabetes, Cardiovascular disease, etc).

**Table 3:**
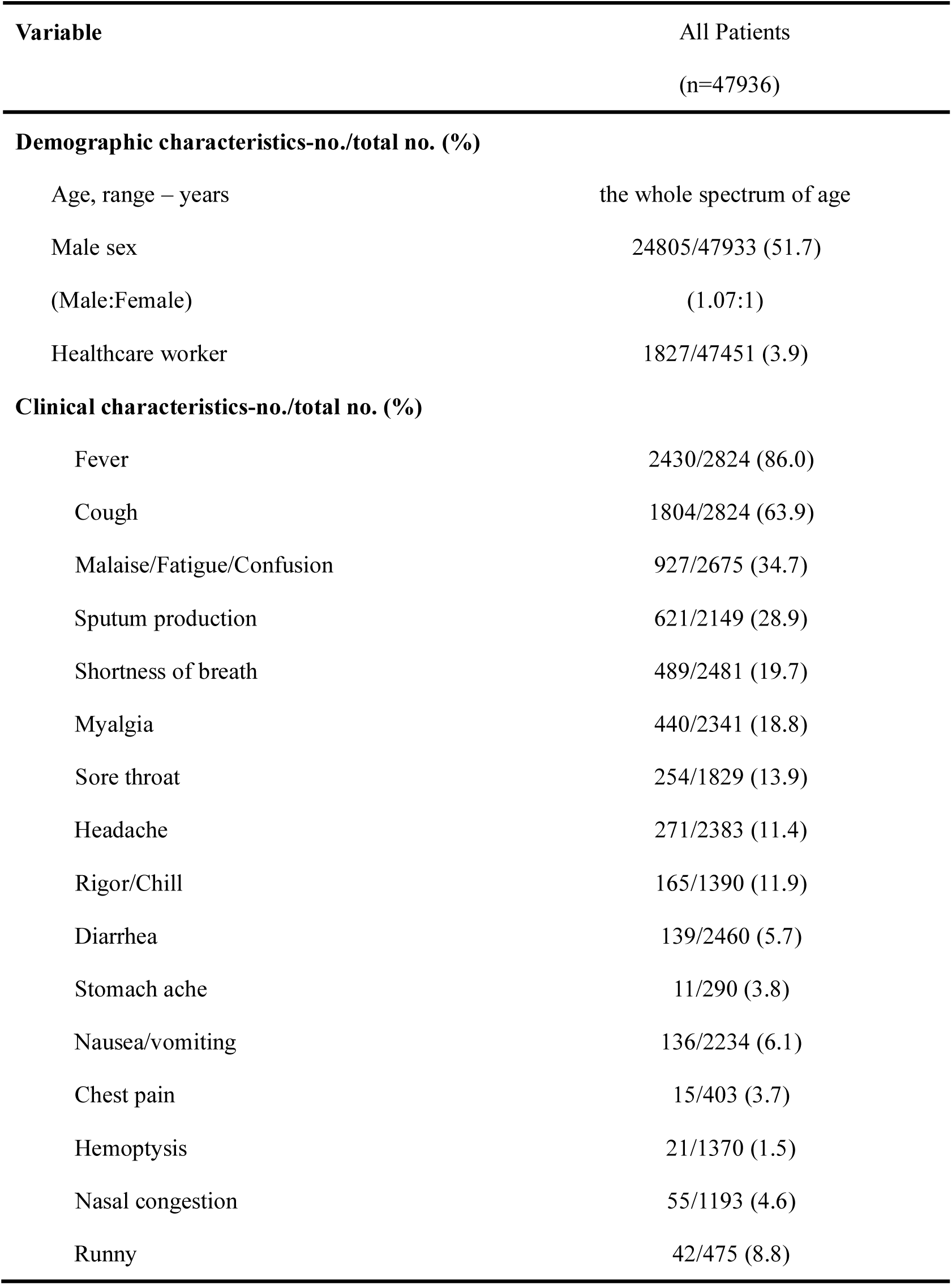
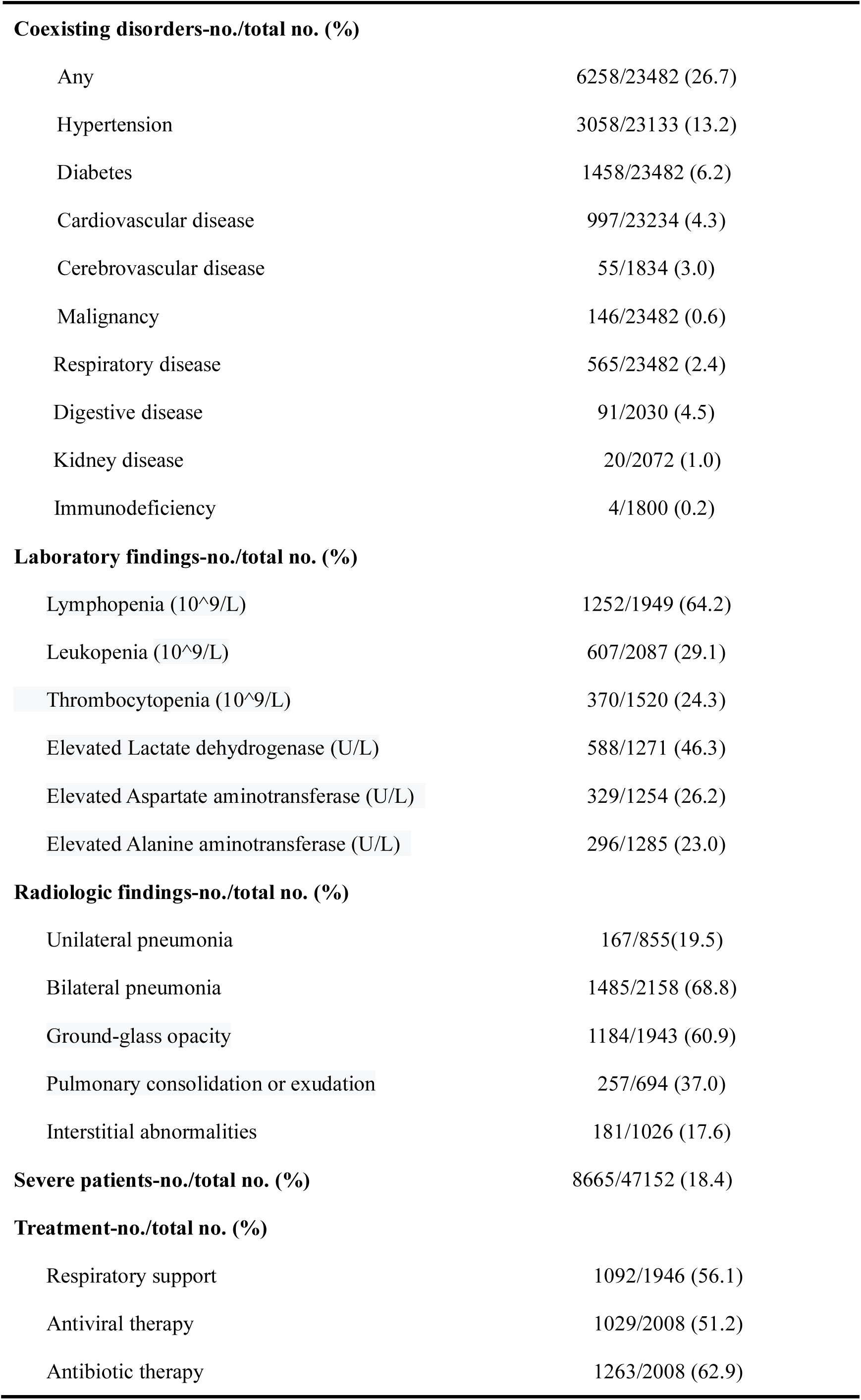
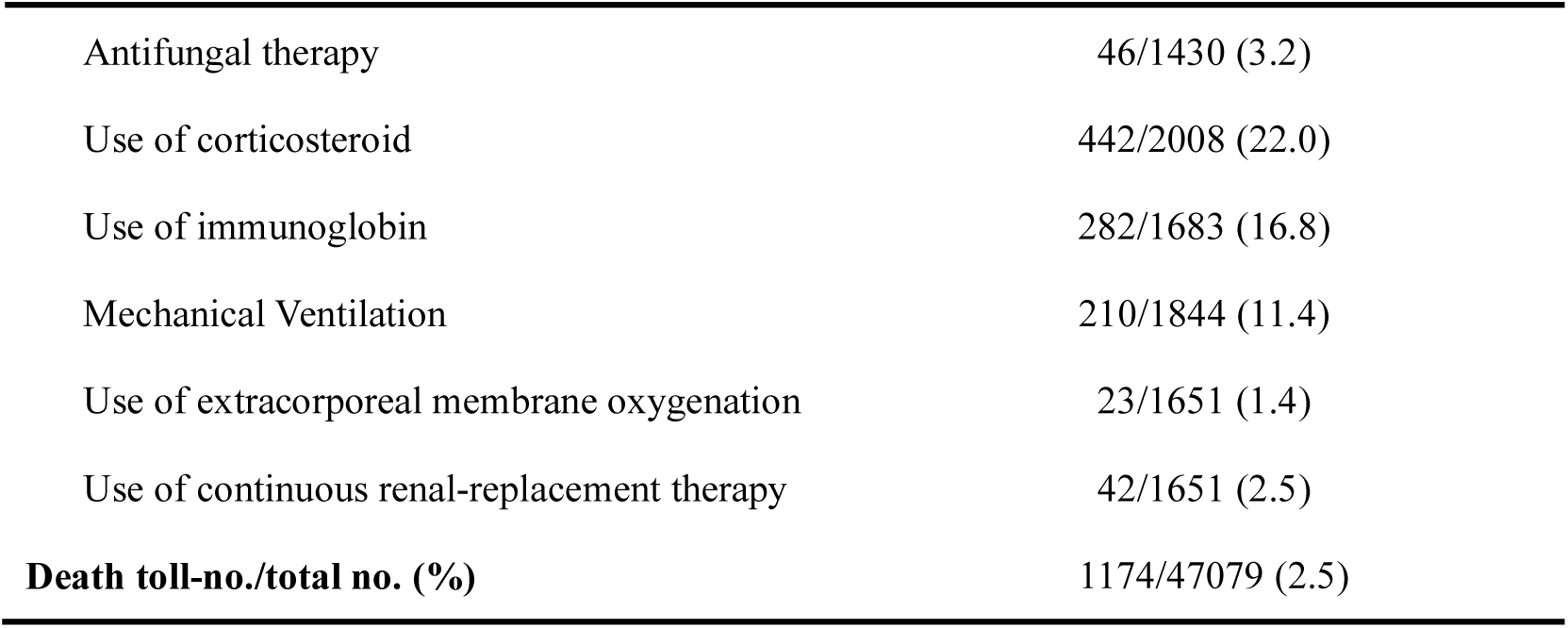
The Summary of Clinical characteristics and auxiliary examination of patients infected with COVID-19

| Variable | All Patients<br>(n=47936) |
| --- | --- |
| **Demographic characteristics-no./total no. (%)** |  |
| Age, range – years | the whole spectrum of age |
| Male sex | 24805/47933 (51.7) |
| (Male:Female) | (1.07:1) |
| Healthcare worker | 1827/47451 (3.9) |
| **Clinical characteristics-no./total no. (%)** |  |
| Fever | 2430/2824 (86.0) |
| Cough | 1804/2824 (63.9) |
| Malaise/Fatigue/Confusion | 927/2675 (34.7) |
| Sputum production | 621/2149 (28.9) |
| Shortness of breath | 489/2481 (19.7) |
| Myalgia | 440/2341 (18.8) |
| Sore throat | 254/1829 (13.9) |
| Headache | 271/2383 (11.4) |
| Rigor/Chill | 165/1390 (11.9) |
| Diarrhea | 139/2460 (5.7) |
| Stomach ache | 11/290 (3.8) |
| Nausea/vomiting | 136/2234 (6.1) |
| Chest pain | 15/403 (3.7) |
| Hemoptysis | 21/1370 (1.5) |
| Nasal congestion | 55/1193 (4.6) |
| Runny | 42/475 (8.8) |
| **Coexisting disorders-no./total no. (%)** |  |
| Any | 6258/23482 (26.7) |
| Hypertension | 3058/23133 (13.2) |
| Diabetes | 1458/23482 (6.2) |
| Cardiovascular disease | 997/23234 (4.3) |
| Cerebrovascular disease | 55/1834 (3.0) |
| Malignancy | 146/23482 (0.6) |
| Respiratory disease | 565/23482 (2.4) |
| Digestive disease | 91/2030 (4.5) |
| Kidney disease | 20/2072 (1.0) |
| Immunodeficiency | 4/1800 (0.2) |
| **Laboratory findings-no./total no. (%)** |  |
| Lymphopenia (10<sup>9</sup>/L) | 1252/1949 (64.2) |
| Leukopenia (10<sup>9</sup>/L) | 607/2087 (29.1) |
| Thrombocytopenia (10<sup>9</sup>/L) | 370/1520 (24.3) |
| Elevated Lactate dehydrogenase (U/L) | 588/1271 (46.3) |
| Elevated Aspartate aminotransferase (U/L) | 329/1254 (26.2) |
| Elevated Alanine aminotransferase (U/L) | 296/1285 (23.0) |
| **Radiologic findings-no./total no. (%)** |  |
| Unilateral pneumonia | 167/855(19.5) |
| Bilateral pneumonia | 1485/2158 (68.8) |
| Ground-glass opacity | 1184/1943 (60.9) |
| Pulmonary consolidation or exudation | 257/694 (37.0) |
| Interstitial abnormalities | 181/1026 (17.6) |
| **Severe patients-no./total no. (%)** | 8665/47152 (18.4) |
| **Treatment-no./total no. (%)** |  |
| Respiratory support | 1092/1946 (56.1) |
| Antiviral therapy | 1029/2008 (51.2) |
| Antibiotic therapy | 1263/2008 (62.9) |
| Antifungal therapy | 46/1430 (3.2) |
| Use of corticosteroid | 442/2008 (22.0) |
| Use of immunoglobulin | 282/1683 (16.8) |
| Mechanical Ventilation | 210/1844 (11.4) |
| Use of extracorporeal membrane oxygenation | 23/1651 (1.4) |
| Use of continuous renal-replacement therapy | 42/1651 (2.5) |
| **Death toll-no./total no. (%)** | **1174/47079 (2.5)** |

### Imaging and laboratory findings at presentation

**Table 3** shows the laboratory and Imaging findings In COVID-19. In these review we found the most common, patterns on chest imaging findings were ground-glass opacity (60.9%) and pulmonary consolidation or exudation (37.0%), 19.5% were unilateral pneumonia and 68.8% were bilateral pneumonia. 64.2%, 29.1% and 24.3% of patients had lymphopenia, leucopenia and thrombocytopenia, respectively.

### Treatment and outcomes

**Table 3** shows the treatment and outcomes. In this review we found that in COVID-19, oxygen therapy, antiviral therapy, antibiotic therapy, the use of corticosteroids or immunoglobulin were in 56.1%, 51.2%, 62.9%, 22.0% and 16.8% of patients, respectively. Another 1.4% and 2.5% of people use of extracorporeal membrane oxygenation and continuous renal-replacement therapy. The death toll and severe patients of COVID-19 were 18.4% and 2.5%.

### Mode of transmission

Before the closes of Huanan seafood market on January 1, 2020. people contracted the disease mainly through contact with host animals carrying the virus^[24]^, Subsequently, infected people spread the virus to their families, communities through human-to-human transmission and contact transmission. Other countries are also seeing increasing numbers of travel-related imported cases and human-to-human transmission.

### Pulmonary pathology

**Table 4** summarizes the pulmonary pathological features of COVID-19. The pathology showed that the manifestations of COVID-19 were similar to SARS. But the latest pathological examination of COVID-19 revealed the obvious mucinous secretions in the lungs.

**Table 4:** Pulmonary pathology of SARS-CoV-2 pneumonia

| Pulmonary pathology | SARS-CoV-2<sup>[41-43]</sup> |
| --- | --- |
| Case number | 4 |
| Pathological manifestations | Bilateral diffuse alveolar injury with dense fibrous mucous exudates. Significant pulmonary oedema and fibrin exudate in alveolar edema. have desquamation of pneumocytes and hyaline membrane formation. There is patchy and severe pulmonary cell proliferation and interstitial thickening with inflammatory cells, multinucleated giant cells and macrophage infiltration. These features associated with acute respiratory distress syndrome. |
| Pathological features | Prominent fibrous mucus exudate and mucous plug formation. |

## Discussion

SARS-CoV and SARS-CoV-2 are both belong to the β coronavirus family, which share 78% of their nucleotide homology^[18]^. As more and more confirmed cases of COVID-19 have been reported, we found that SARS and COVID-19 have great similarities in clinical manifestations, laboratory examinations and chest imaging.

### Infectious source

Current studies have shown that SARS-CoV-2 is mainly related to wildlife trafficking in Huanan seafood market in China. A study by Chan et al found that SARS-CoV-2 is most closely related to coronaviruses found in horseshoe bats in China^[44]^.The SARS outbreak in 2002 and the Middle East respiratory syndrome (MERS) outbreak in 2012 have demonstrated the possibility of coronavirus transmission from animals to humans[45, 46].

It is currently believed that whether SARS or COVID-19, the main source of infection is people infected with coronavirus. in the early report of 41 confirmed cases of COVID-19, 27 reported exposure to the Huanan seafood market^[24]^. But since the closure of the seafood market on Jan 1, 2020, more and more new cases have been reported to be associated with contact with confirmed patients which stress the importance of human-to-human transmission. However, it’s worth noting that there were some people uninfected in close contacts with SARS or COVID-19^[12, 13, 16]^. Therefore, whether genetic susceptibility exists or not needs further study.

### Route of transmission

In this study, we found COVID-19 were mainly transmitted by droplets and contact, showing human-to-human transmission, family aggregation spread and nosocomial infection. It is worth noting that COVID-19 has a more diverse mode of transmission that can manifest itself as asymptomatic infection^[7]^, something that has not previously been seen in SARS^[47]^. Because of the absence of symptoms, it is difficult to detect and isolate carriers in time, which makes it more difficult to control the spread of the disease^[47, 48]^. Recent studies have found that SARS-CoV-2 can be detected in the feces of patients, indicating that the possibility of fecal-oral transmission^[4, 12]^. There have been reports of infections in newborns^[2]^, but more studies are needed to confirm the presence of vertical transmission. A study of nine pregnant women^[2]^ which tested the amniotic fluid, cord blood, neonatal throat swabs and breast milk of six patients for SARS-CoV, all of which were negative and did not support mother-to-child transmission. Human-to-human, asymptomatic infection and possible mother-to-child transmission and fecal-oral transmission are the causes of the widespread spread of the disease. So, the early recognition of COVID-19 in different countries can reduces transmission risk and increases understanding of SARS-CoV-2, to inform national and global response actions.

### Susceptible population

Through the summary analysis of the pneumonia, we found that COVID-19 had a general susceptibility, However, it should be noted that cases^[2]^ of infection in pregnant women, newborns, infants and children have been reported successively. The reason was considered to be related to the special immune tolerance state of pregnant women during pregnancy^[2]^ and the low immune function of children and infants^[48]^. Elderly people with basic diseases are at high risk of infection with COVID-19,due to the low body defense against infection, it is easy to develop severe pneumonia once infected with COVID-19^[12, 21]^.

### Clinical manifestations, auxiliary examination and treatment

Through the study of COVID-19 in different countries, we found that pneumonia were mainly characterized by fever, accompanied by different degrees of cough, dyspnea, fatigue and discomfort. However, the clinical symptoms of COVID-19 were not obvious. COVID-19 patients with mild symptoms are easy to be overlooked, so it is necessary to take active and effective isolation measures for suspected patients or close contacts as early as possible. Furthermore, laboratory tests showed that patients with COVID-19 may had lymphopenia, leukopenia, and thrombocytopenia, along with elevated levels of liver enzymes and lactate dehydrogenase.

On imaging, COVID-19 was mainly manifested as multiple ground-glass shadows. A CT scan of 29 patients with COVID-19 revealed a single or multiple patchy frosted glass shadow with septum thickening in typical case of COVID-19.As the disease progresses, the lesion increases and the scope expands, and frosted glass shadows coexist with solid or striated shadows^[29]^. An imaging study of 81 people found that abnormal chest imaging could occur even in asymptomatic patients and progressed rapidly from focal unilateral to diffuse bilateral within 1-3 weeks^[26]^. Since the early imaging changes of COVID-19 were not obvious, high resolution chest CT examination should be performed as soon as possible for suspected patients to clarify. At present, the treatment of COVID-19 is lack of specific drugs and mainly focuses on support treatment. Therefore, early detection, early isolation, early diagnosis and early treatment are the key measures to control infectious diseases. Our clinical and pathological review of COVID-19 can help healthcare worker to formulate a timely therapeutic strategy for similar patients and reduce mortality

### Pathological characteristics

SARS-CoV-2 has been identified as a sister virus to SARS-CoV, a SARS-related coronavirus (SARSr-CoV). SARS-CoV and SARS-CoV-2 have been shown to infect human respiratory epithelial cells through the interaction of viral S proteins and angiotensin-converting enzyme 2 receptors on human cells^[26, 49]^. Therefore, it is speculated that they may have the same pathological changes. The early pathology of SARS found^[50, 51]^ that SARS-CoV widespreadly existed in the epithelium of the digestive tract, the epithelium of the distal convoluted tubules of the kidney, and the sweat glands of the skin, suggesting that in addition to respiratory transmission, SARS might also be transmitted through contact between the patient’s feces, urine and skin. which also provided a new way to prevent the transmission of COVID-19. In addition, Ding et al^[50, 51]^ also found SARS-CoV in systemic endocrine gland, Because SARS-CoV and SARS-CoV-2 have the same mechanism of action, they can cause rapid production of multiple cytokines in body fluids after infection with microorganisms, leading to acute respiratory distress and multiple organ failure. This also explains why most covid-19 patients have mild symptoms at the onset of the disease, while some are suddenly worse after being diagnosed in hospital, which may be related to the body producing too many cytokines after the disease, leading to “cytokine storm” in the body.

Xu et al^[42]^ and Tian et al^[41]^ found diffuse alveolar injury, transparent membrane formation and pulmonary edema in the early pulmonary puncture pathology of COVID-19 patients, which was similar to the pathological manifestations of SARS. However, it is worth noting that the pathology of COVID-19 also shows fibromyxoid exudation and the formation of thick “mucus plugs”. The first autopsy, performed by liu’s team^[43]^, found a large amount of sticky secretions spilling out of the alveoli, with fibrous cords visible. As of February 25, Liu’s team completed a preliminary anatomical diagnosis of three of the 11 cases, all of which found mucinous secretions in the lungs of the deceased. This finding is a warning for clinical treatment. If the mucus components are not dissolved, oxygen alone may not achieve the goal, and sometimes may be counterproductive to increase the hypoxia of patients.

#### Limitations

This study has several limitations, first, among those cases, most patients are still hospitalized at the time of paper published. Therefore, it is difficult to accurate assess number of deaths and severe cases, and risk factors for poor outcome. so continued observations of the process and prognosis of the disease are needed. second,So far, many foreign reports on COVID-19 have been individual cases,making comparisons difficult. This paper mainly uses descriptive analysis to review and summarize the case.

##### Conclusion

The source, intermediate host, transmission route and incubation period of SARS-CoV-2 are not fully understood. Knowledge of the virus remains elusive and unpredictable. Therefore, early pathological examination is the most direct means to uncover the true face of SARS-CoV-2. Now the outbreak of COVID-19 constitutes an epidemic threat in China and Worldwide. It is important for public medical institutions, health care workers, and the public to have an early recognition of COVID-19 so that coordinated and effective actions can help prevent additional cases or poor health outcomes.

## Data Availability

The data used to support the findings of this study are included within the article.

## Acknowledgements

We greatly appreciate the efforts of all the hospital employees and their families at the Fujian Provincial Hospital and affiliated Hospital of Fujian Medical University, who are working tirelessly during this outbreak.

## Article Information

### Author Contributions

Gang Chen had full access to all of the data in the study and takes responsibility for the integrity of the data and the accuracy of the data analysis.

### Concept and design

Yaqian Mao, Wei Lin,, Gang Chen.

### Acquisition, analysis, or interpretation of data

Yaqian Mao,Wei Lin, Junping Wen

### Drafting of the manuscript

Yaqian Mao, Gang Chen

### Critical revision of the manuscript for important intellectual content

Yaqian Mao, Gang Chen

### Statistical analysis

Yaqian Mao

### Administrative, technical, or material support

None

### Supervision

Gang Chen

